# Evidence for Aerosol Transfer of SARS-CoV2-specific Humoral Immunity

**DOI:** 10.1101/2022.04.28.22274443

**Authors:** Ross M. Kedl, Elena Hsieh, Thomas E. Morrison, Gabriela Samayoa-Reyes, Siobhan Flaherty, Conner L. Jackson, Rosemary Rochford

## Abstract

Despite the obvious knowledge that infectious particles can be shared through respiration, whether other constituents of the nasal/oral fluids can be passed between hosts has surprisingly never even been postulated, let alone investigated. The circumstances of the present pandemic facilitated a unique opportunity to fully examine this provocative idea. The data we show provides evidence for a new mechanism by which herd immunity may be manifested, the aerosol transfer of antibodies between immune and non-immune hosts.

## Introduction

The vaccines against SARS-CoV-2 have maintained remarkable efficacy against severe disease and death in those vaccinated regardless of variant emergence, Omicron included ^1^. Less appreciated than the systemic immunity generated by the vaccines are the high levels of antibody (IgG and IgA) found within the nasal cavity and saliva of vaccinees. This outcome is found in both humans and primates, and in response to both mRNA and protein-based vaccines ^2,3^. Respiratory transmission of viral infection is proof that oral/nasal cavity constituents can be communicated through aerosols and/or respiratory droplets. As such, it would stand to reason that antibody present within the oral/nasal environment may also be aerosolized to some degree.

## Results

The extended mandates for mask wearing in both social and work environments provided a unique opportunity to evaluate the possibility of aerosolized antibody expiration from vaccinated individuals. Utilizing a flow cytometry-based Multiplex Microsphere Immunoassay (MMIA) to detect SARS-CoV-2-specific antibodies (**Fig 1A and B**) ^4,5^ and a method previously used to elute antibody from rehydrated dried blood spots (DBS), we identified anti-SARS-CoV-2 specific antibodies eluted from surgical face masks worn by vaccinated lab members donated at the end of one workday. Consistent with the results reported by others, we identified both IgG and IgA in saliva from vaccinated individuals (**Fig 1C and D**). It was therefore not surprising to detect both IgG and IgA following elution of antibody from face masks (**Fig 1C and D**).

**Figure 1.**
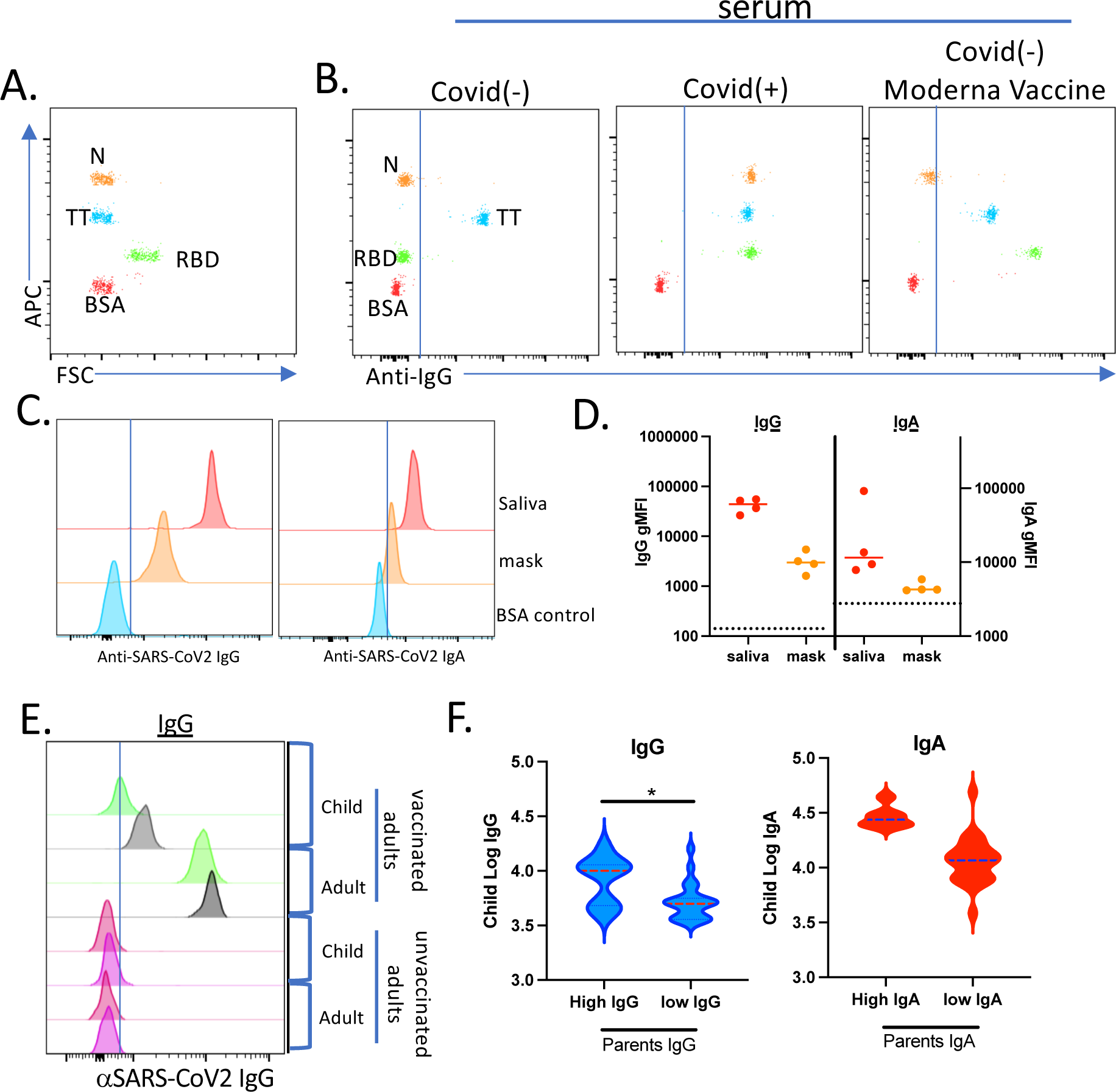
Evidence for aerosol transfer of SARS-CoV2-specific immunity. A and B. representative flow cytometric results from the use of a Multiplex Microsphere Immunoassay (MMIA) (A) evaluating serum samples from a Covid(-) (B, left), Covid(+) (B, middle), and Moderna mRNA vaccinee (B, right). C and D. Histograms showing the mean fluorescent intensities for Wuhan-RBD-specific IgG (left) and IgA (right) from saliva or eluted from a surgical mask worn for one workday. D. Quantification of IgG and IgA gMFI eluted from masks obtained from 4 individuals. Dotted lines indicate gMFI obtained for Covid/vaccine (-) sample. E. Histograms showing the mean fluorescent intensities for Wuhan-RBD-specific IgG eluted from nasal swabs from unvaccinated children living in households in which parents or family members were either vaccinated (top) or unvaccinated (bottom). F. Log transformation of the gMFI for Wuhan-RBD-specific IgG (left) or IgA (right) from thirty-four adult-child pairs using antibody cut-offs for hi vs low parental intranasal antibody levels.

Given these observations, we hypothesized that droplet/aerosolized antibody transfer might occur between individuals, much like droplet/aerosolized virus particles can be exchanged by the same route. To evaluate this hypothesis, we obtained nasal swabs from children living in households in which parents or family members had varying degrees of SARS-CoV2-specifc immunity, including those unvaccinated, vaccinated and COVID-19+. Initial comparison of nasal swabs acquired from children living in vaccinated households revealed readily detectable SARS-CoV-2-specific IgG (**Fig 1E**), especially when compared to the complete deficit of SARS-CoV-2-specific antibody detected in the few nasal swabs we obtained from children in non-vaccinated households. We then used the variation in parents’ levels of intranasal IgG as the basis of stratification across all children’s samples. Log transformation of the data from thirty-four adult-child pairs established antibody cut-offs for high vs low parental intranasal antibody levels. Evaluation of samples in this fashion revealed that high intranasal IgG in vaccinated parents was significantly associated (p-value = 0.01) with a 0.38 increase in the log transformed intranasal IgG gMFIs within a child from the same household (**Fig 1F**). This significant positive relationship was observed using either parametric or non-parametric analysis, and adjustments for the correlation within household did not alter the conclusion. Though not statistically significant, a similar trend of elevated IgA was found in the same samples.

## Discussion

The concept of herd immunity is a central tenant of public health vaccination campaigns. Overt blockade of infection as well as a reduction in viral transmission downstream of a breakthrough infection are widely accepted conceptual mechanisms by which vaccination-induced immunity in specific individuals protects non-immune community members. Our results suggest that aerosol transmission of antibodies may also contribute to host protection and represent an entirely unrecognized mechanism by which passive immune protection may be communicated. Whether antibody transfer mediates host protection will be a function of exposure, but it seems reasonable to suggest, all things being equal, that any amount of antibody transfer would prove useful to the recipient host. A recent publication showed substantial benefits of parental vaccination in reducing the risk of infection in the unvaccinated children in the same home ^6^. It is tempting to speculate that aerosol-mediated antibody transfer may have possibly contributed to their findings reported.

## Methods

A Multiplex Microsphere Immunoassay (MMIA) was constructed and performed as previously described ^4^. Under IRB # 20-1279, serum samples were obtained from first-responder adults in Arapahoe County, CO ^5^. Antibody was eluted ^4^ from punches taken from the center of surgical masks anonymously donated by laboratory workers. Nasal swabs were obtained by convenience sampling both parents and their children at the Colorado Tricountry vaccine center in Aurora, CO who were attending vaccine appointments, not limited to SARS-CoV2. Antibody from swab tips was eluted as described for DBS ^4^. The log transformed IgA and IgG values from the children’s samples were modeled using linear regression with a single binary covariate corresponding to high or low antibody levels from their parent. Residual plots were used to check violations of linear regression assumptions and a Wilcoxon rank sum test was conducted if assumptions were violated. A linear mixed effects model was evaluated to assure that the correlation within household did not significantly contribute to the data or alter the conclusions drawn from the fixed effect linear regression model. Cytometry was performed using a Beckman Coulter Cytoflex cytometer and analyzed using FloJo v.10 software (Treestar, Inc.). Statistical analyses were conducted using R (version 4.0.2).

## Data Availability

All materials, data, and associated protocols will be available to readers upon request and without undue qualifications

## Funding Statement

Funding for these studies came from Academic Enrichment Funds (RMK, RR and, and TEM) and was not provided through any grant or agency.

## Notes

Declarations: The authors have no conflict of interests to declare.

### Competing Interest Statement

The authors have declared no competing interest.

### Funding Statement

This study did not receive any funding

### Author Declarations

Acquisition of clinical samples was performed in accordance with approval from the Institutional Review Board at the University of Colorado Anschutz Medical campus, protocol #20-1279.

